# Age prediction using resting-state functional MRI

**DOI:** 10.1101/2023.12.26.23300530

**Authors:** Jose Ramon Chang, Zai-Fu Yao, Shulan Hsieh, Torbjörn E. M. Nordling

## Abstract

The increasing lifespan and large individual differences in cognitive capability highlight the importance of comprehending the aging process of the brain. Contrary to visible signs of bodily ageing, like greying of hair and loss of muscle mass, the internal changes that occur within our brains remain less apparent until they impair function. Brain age, distinct from chronological age, reflects our brain’s health status and may deviate from our actual chronological age. Notably, brain age has been associated with mortality and depression. The brain is plastic and can compensate even for severe structural damage by rewiring. Functional characterization offers insights that structural cannot provide. Contrary to the multitude of studies relying on structural magnetic resonance imaging (MRI), we utilize resting-state functional MRI (rsfMRI). We also address the issue of inclusion of subjects with abnormal brain ageing through outlier removal.

In this study, we employ the Least Absolute Shrinkage and Selection Operator (LASSO) to identify the 39 most predictive correlations derived from the rsfMRI data. The data is from a cohort of 176 healthy right-handed volunteers, aged 18-18 years (9 81 male female, mean age 8, SD 11) collected at the Mind Research Imaging Center at the National Cheng Kung University.

We establish a normal reference model by excluding 68 outliers, which achieves a leave-one-out mean absolute error of 2. 8 years. By asking which additional features that are needed to predict the chronological age of the outliers with a smaller error, we identify correlations predictive of abnormal aging. These are associated with the Default Mode Network (DMN).

Our normal reference model has the lowest prediction error among published models evaluated on adult subjects of almost all ages and is thus a candidate for screening for abnormal brain aging that has not yet manifested in cognitive decline. This study advances our ability to predict brain aging and provides insights into potential biomarkers for assessing brain age, suggesting that the role of DMN in brain aging should be studied further.

## 1 Introduction

The ageing process is a complex and dynamic phenomenon, marked by progressive physiological and functional changes that occur throughout an individual’s lifespan (Sinclair and Oberdoerffer, 2009). While chronological age serves as a fundamental indicator of the passage of time, it often falls short of capturing the true age-related alterations within the human brain. Individuals of the same chronological age can exhibit distinct patterns of cognitive decline or preservation, indicating the existence of an intriguing phenomenon known as the “brain age gap” (Ballester et al., 2023; Sanford et al., 2022; Jawinski et al., 2022; Niu et al., 2020; Mohajer et al., 2020).

The brain age gap refers to the discrepancy between the biological age of an individual, as inferred from the structural and functional characteristics of their brain, and their chronological age. Advancements in neuroimaging techniques, coupled with cutting-edge machine learning algorithms, have opened up unprecedented opportunities to quantify this age gap and explore its implications for human health and cognition (Baecker et al., 2021; Lee et al., 2022a). In recent years, the investigation of brain age gaps has garnered significant attention in neuroscience and aging research. Understanding why and how the biological age of the brain can differ from the chronological age holds the promise of unveiling new insights into the mechanisms that govern brain aging and cognitive decline (Lee et al., 2022b; Elliott et al., 2021). Additionally, studying brain age gaps provides invaluable tools for assessing individual health trajectories, identifying neurodegenerative risk factors, and tailoring personalized interventions to promote healthy ageing (Franke and Gaser, 2019; Ran et al., 2022).

Analysis involving rsfMRI often links abnormal aging to the DMN, given its activity during periods of rest and when the mind is not focused on the external world (Wang et al., 2020). It is also well-established that patients with Alzheimer’s disease exhibit reduced functional connectivity within the DMN (Ibrahim et al., 2021; Kucikova et al., 2021). However, other brain networks have garnered attention as pertinent to age prediction. Podg6rski et al. (2021) reported that in the process of brain aging in elderly females, various networks, including the visual, salience, sensorimotor, language, frontoparietal, dorsal attention, and cerebellar networks, compensate for these changes. Conversely, Oschmann et al. (2020) found that during longitudinal observations of subjects, changes were evident in the frontoparietal and salience networks but not in the DMN. Additionally, Liu et al. (2022) asserted that the attention and frontoparietal networks are more relevant to aging than the DMN. Our objective is not only to create a model capable of predicting brain age but also to identify the regions associated with abnormal ageing.

### 1.1 Statistical methods to determine brain age

Table 1 and Table 2 show different statistical models used to predict chronological age from neuroimages. Table 1 shows the methods utilized for each model and Table 2 their respective performances. Studies that did not present a mathematical model, utilized neuroimages, or did not focus on chronological age prediction were excluded.

**Table 1:**
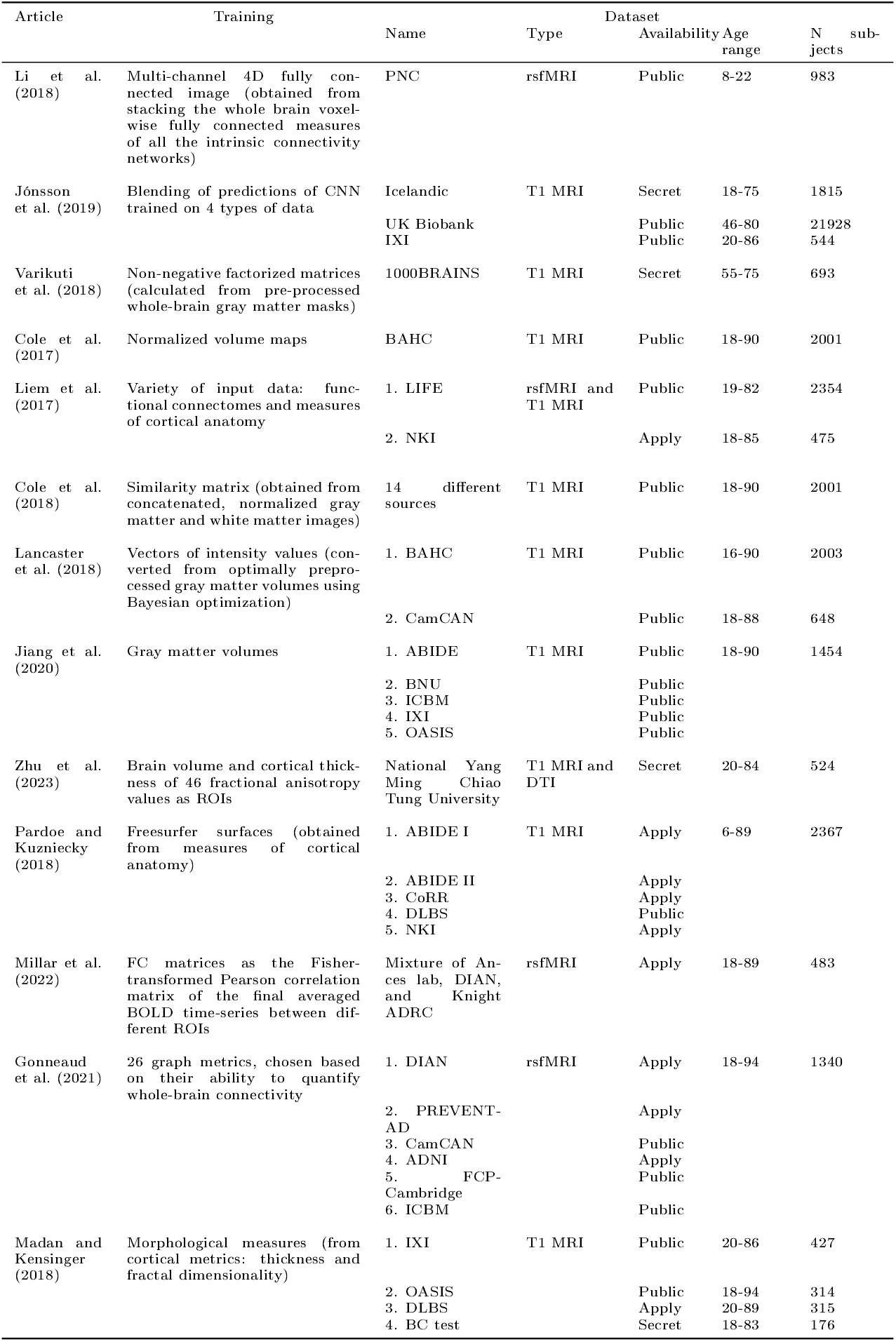
Methods and data sets used for prediction of chronological age from brain images.

**Table 2:** Models and their performance for prediction of chronological age from brain images. Only the best results are presented. Abbreviations: Convolutional Neural Network (CNN), Relevance Yector Machine (RVM), Gaussian Process Regression (GPR), Support Yector Machine (SVM), Artificial Neural Network (ANN), Support Yector Regression (SVR), Correlation Based Regression (CBR), Cross-Yalidation (CV).

| Article | Model | Validation | Performance |  |  |  |
| --- | --- | --- | --- | --- | --- | --- |
| | | | MAE | RMSE | Pearson's<br>r | $R^2$ |
| Li et al.<br>(2018) | CNN | 5-fold CV | 2.15 |  | 0.614 |  |
| Jónsson<br>et al.<br>(2019) | CNN | Split into train (1171),<br>validation (298) and test<br>(346) sets | 3.39 |  |  | 0.87 |
|  |  | 10-fold CV | 3.85 |  |  | 0.77 |
| Varikuti<br>et al.<br>(2018) | LASSO | 3-fold CV | 3.4 |  |  |  |
| Cole<br>et al.<br>(2017) | CNN | Split train (1601), val-<br>idation (200), & test<br>(200) sets | 4.16 | 5.31 | 0.96 | 0.92 |
| Liem<br>et al.<br>(2017) | SVR | Tested on independent<br>data set (NKI) | 4.28 |  |  | 0.87 |
| Cole<br>et al.<br>(2018) | GPR | 10-fold CV | 5.02 | 6.31 | 0.94 | 0.88 |
| Lancaster<br>et al.<br>(2018) | SVM | Tested on independent<br>data set (CamCAN) | 5.08 |  | 0.941 | 0.89 |
| Jiang<br>et al.<br>(2020) | CNN | Split into train (1303)<br>and test (151) sets | 5.55 |  |  |  |
| Zhu<br>et al.<br>(2023) | GPR | Split into train (330) and<br>test (194) sets | 5.56 |  |  |  |
| Pardoe<br>and<br>Kuzniecky<br>(2018) | RVM | Split into train (2167) &<br>test (200) sets | 7.2 |  |  |  |
| Millar<br>et al.<br>(2022) | GPR | Split into train (391) and<br>test (98) sets | 8.20 | 10.32 |  | 0.73 |
| Gonneaud<br>et al.<br>(2021) | SVM | Split into train (773),<br>validation (46), and test<br>(521) sets | 11.58 | 13.24 |  | 0.36 |
| Madan<br>and<br>Kensinger<br>(2018) | RVM | Split into train (1056) &<br>test (176) sets |  |  |  | 0.80 |

It is important to note that the age ranges vary in all studies. Researchers have concluded that performances tend to be better with smaller age ranges (de Lange et al., 2022). Performance metrics used for evaluating age prediction models depend on cohort and study-specific data characteristics, and this needs to be considered when comparing them across different studies. This can also be observed in the studies we considered. The best-performing model (Li et al., 2018), with a Mean Absolute Error (MAE) between predicted brain age and chronological age of 2.15 years, only had an age range of s to 22 years. A sufficient age range is crucial because a naive model that predicts the age of each subject to be the mean of the age range (15 years) has an expected MAE of 3.1 years for the age distribution reported in (Satterthwaite et al., 2014), from where Li et al. (2018) took 983 of the 1445 samples. Any model performing worse than this naive model is arguably worthless. The MAE achieved by Li et al. (2018) is statistically significantly better than this naive model (p-val *<* 0.0001), but if they had selected the age range 11 to 19, then a MAE below 2.15 years would have been seen in 17% of random subject choices with replacement from their age distribution. All models with an age range covering essentially the life span of a human have reported MAEs above 3.39 years.

We have observed that the most popular type of neuroimage is T1 structural MRI, followed by resting-state functional MRI. The most widely used measure of the performance of models is the MAE. This metric is defined as the average absolute value of the residuals between the predicted age and the chronological age across a set of samples. Being the average prediction error makes it easy to understand. The most common method to evaluate the performance of brain age prediction models is to use a separate data set reserved for testing of performance. Cross-validation is also common.

Additionally, we would like to note that most of the efforts focused on age prediction have been cross-sectional studies. We found only one study (Aamodt et al., 2023) that focused on longitudinal age prediction at 18 and 36 months on a cohort of Norwegian subjects. They trained an XGBoost regression model but only reported their mean baseline Brain Age Gap (BAG), which is the residual of the real age minus the predicted age, as 0.18 with a standard deviation of 9.03. Unlike the Mean Absolute Error, BAG does not take the absolute value of the residuals before calculating the mean, thus making the metrics not comparable.

All works on brain age prediction use chronological age as ground truth for training and validation. Since changes to the brain, e.g. in Alzheimer’s disease (Bateman et al., 2012; Preische et al., 2019), occur before the function is visibly impaired, this use of chronological age as ground truth is problematic. It deserves more attention than it has gotten so far in the literature. Moreover, mental, neurological, and substance use disorders constitute around 25% of years lived with disability and the incidence of mental and neurological disorders vary across age groups (WHO, 2023; Nichols et al., 2022; Kang et al., 2022). Thus even a carefully vetted dataset of healthy subjects is likely to include subjects where the brain is not ageing normally. These subjects will bias the data and predictions of a model trained on it in an unknown way that is likely to be highly dependent on the dataset in question. Since this inclusion of subjects with abnormally ageing brains in practice is unavoidable we here explore the usage of outlier removal to exclude these subjects and obtain a model of normal healthy ageing.

## 2 Materials and Methods

### 2.1 Participants

Out of the 205 participants, 15 were excluded due to reasons such as not completing the experiment, not meeting the MRI criteria, etc. The participant demographic information is shown in Table 3. All participants were assessed with the Montreal Cognitive Assessment (MoCA) (Nasreddine et al., 2005) and Beck Depression Inventory-II (BDI) (Beck et al., 1996). A total of 11 participants with scores lower than 22 on the MoCA (n=4) and higher than 13 on the BDI-II (n=7) were excluded during the data analysis. Additionally, 3 individuals were not included in the analysis due to image quality issues. Out of the remaining 176 subjects healthy right-handed volunteers aged 18-78 years (81 females, mean age 48.04, std = 16.81), 156 had their data recorded prior to the MRI software update, while the remaining 20 were enrolled after the update.

**Table 3:** Demographic information of the 176 participants.

| Age group<br>(years) | Age<br>(mean/ std) | Gender<br>(male/female) | Education<br>years<br>(mean/std) | MoCa<br>(mean/std) | Cardiovascular<br>risk factor<br>(+/-) | Before MRI<br>update<br>(before/after) |
| --- | --- | --- | --- | --- | --- | --- |
| 18-30 | 24.24/2.95 | 25/15 | 16.05/1.52 | 28.73/0.95 | 14/25 | 34/6 |
| 30-40 | 34.00/2.93 | 10/9 | 15.95/1.54 | 27.47/1.93 | 9/10 | 18/1 |
| 40-50 | 44.94/2.91 | 15/10 | 14.24/2.3 | 26.64/2.19 | 10/15 | 24/1 |
| 50-60 | 55.30/3.04 | 14/23 | 14.38/2.69 | 26.97/1.88 | 23/14 | 35/2 |
| 60-70 | 64.69/2.35 | 21/21 | 14.10/3.01 | 27.29/1.78 | 21/21 | 32/10 |
| 70-80 | 73.29/2.35 | 10/3 | 13.92/2.62 | 26.23/1.89 | 3/10 | 13/0 |

In addition, other metadata such as the gender, education years, and cardiovascular risk factors of the subjects were recorded. Cardiovascular risk factors include diabetes, hypertension, hyperlipidemia, hyperglycemia, stroke, smoking (past/present), and other reported untreated risk factors. Procedures were carried out in accordance with ethical approval obtained from the National Cheng Kung University Research Ethics Committee, and participants provided written, informed consent before the start of the experiment.

### 2.2 Image acquisition

MRI images were acquired using a GE MR750 3T scanner (GE Healthcare, Waukesha, WI, USA) at the Mind Research Imaging Center at National Cheng Kung University. Resting-state functional images were acquired with a gradient-echo Echo-Planar Imaging (EPI) pulse sequence (TR = 2000 ms, TE = 30 ms, flip angle = 77°, 64 × 64 matrices, FOV = 22 × 22 cm^2^, slice thickness = 4 mm, no gap, voxel size = 3.4375 mm × 3.4375 mm × 4 mm, 32 axial slices covering the entire brain). A total of 245 volumes were acquired; the first five served as dummy scans and were discarded to avoid T1 equilibrium effects. During the scans, participants were instructed to remain awake with their eyes open and fixate on a central white cross displayed on the screen. High-resolution anatomical T1 images were acquired using fast-SPGR, consisting of 166 axial slices (TR = 7.6 ms, TE = 3.3 ms, flip angle = 12°, 224 A−224 matrices, slice thickness = 1 mm), which lasted 218 seconds.

### 2.3 Image processing

For functional connectivity analysis, first, EPI data were pre-processed using the Data Processing and Analysis for Brain Imaging toolbox (Yan et al., 2016), implemented in Matlab (The MathWorks, Inc., Natick, MA, USA), which called functions from SPM 8. Images were slice-time corrected and realigned to correct for head motion using a rigid-body transformation. The mean EPI image was coregistered with the T1 image, and the T1 image was coregistered and normalized to the Montreal Neurological Institute and Hospital (MNI) template. The co-registration parameters of the mean EPI were applied to all functional volumes. Nuisance time series (motion parameters, ventricle, and white matter signals) were regressed out. The functional data were then spatially smoothed with a 6 mm Gaussian kernel. Finally, the images were band-pass filtered at 0.01 to 0.08 Hz to remove scanner drift and high-frequency noise (e.g., respiratory and cardiac activity).

#### 2.3.1 Rest state data preprocessing

Each subject contributed time series data from one resting-state fMRI session. The workflow of functional data preprocessing is summarized as follows: (1) removal of the first four volumes of the Blood-Oxygen-Level Dependent (BOLD) signals to achieve signal stabilization; (2) realignment of functional images using Motion Correlation FMRIB’s Linear Image Registration Tool (MCFLIRT) (Jenkinson et al., 2002); (3) removal of nine confounding signals (six motion parameters plus global, white matter, cerebral spinal fluid) as well as the temporal derivative, quadratic term, and temporal derivatives of each quadratic term (36 regressors in total) (Satterthwaite et al., 2016); (4) co-registration of functional images with the T1 image using boundary-based registration (Greve and Fischl, 2009); (5) alignment of the co-registered images to the template space using the ANTs-transform for the T1 image, as mentioned above; and (6) temporal filtering of time series between 0.01 and 0.08 Hz, as in previous studies (Biswal et al., 1995), using a first-order Butterworth filter. In this study, all regressors, including motion parameters and confound time courses, were band-pass filtered to the same frequency range as the time series data to prevent frequency-dependent mismatch during confound regression (Hallquist et al., 2013). Functional images were smoothed using Gaussian convolution with a 6 mm full-width at half-maximum.

#### 2.3.2 Parcellation

We partitioned the brain of each participant into cortical and subcortical Regions of Interest (ROI)s using the following Power et al. (2011) whole-brain functional atlases (*i*.*e*. 264 cortical and subcortical ROIs of the widely-used functional Power atlas (Power et al., 2011).

#### 2.3.3 Functional connectivity

For each participant, whole-brain functional connectivity between all brain regions was constructed pairwise from the preprocessed fMRI data. The fMRI time series were extracted from each voxel and averaged within each ROI of the three atlases (AAL, Power, and Gordon). The functional connectivity between time series for all pairwise ROIs was estimated by calculating two commonly used connectivity metrics: Pearson’s correlation and wavelet coherence. For Pearson’s correlation, the correlation coefficients were Fisher-transformed to enable drawing more statistically interpretable conclusions about the magnitude of the correlations (Cohen and D’Esposito, 2016; Doucet et al., 2017).

### 2.4 Feature selection and age prediction

Feature selection and age prediction were performed using the LASSO (Tibshirani, 1996). LASSO finds the model parameters that minimise the sum of squared residuals and the sum of absolute parameter values. Given the small number of samples, we aim to create the simplest model that can explain the data. To find the simplest model, i.e. the model with the fewest parameters, we would ideally like to minimise the *L*_0_ norm of the parameters. *L*_0_ is a variation of *L*_*P*_ regularization that penalizes parameters for being different from zero. However, for any *P <* 1 the function describing the parameter value vs. the penalty applied is not convex, and the optimization in general an untractable combinatorial NP-Hard problem. To get around this, we resort to *L*_1_ regularization, i.e. LASSO, which in practice has been shown to yield sparse and predictive models (James et al., 2013). A more detailed explanation of feature selection using LASSO can be found in the Appendix.

The Fisher’s z-transform of Pearson’s r representation of the fMRI’s Functional Connectivity (FC) is a 264 Ã-264 symmetric matrix *A*, with the diagonal containing extremely high values approaching infinity. Since the matrix is symmetric, for any element *a*_*i,j*_ in *A* where *i, j* = 1, 2, …, 264 there exists an identical element *a*_*j,i*_. To eliminate redundant and impractical features, we used either the flattened upper or lower triangle of the matrix *A*, excluding the diagonal, as the input vector of our linear regression model. After pre-processing the data of *n* = 176 subjects, the resulting feature vectors span over a *p* = 34716 dimensional space. This then calls for identifying a subset of *q* ≪ *p* relevant features, also known as regressors, useful for explaining/predicting the target variable *y*, i.e. age. In other words, we need to solve a feature selection problem.

The Glmnet implementation of LASSO enables an elasticnet solution by manipulating the mixing parameter *α*. With range *α* ∈ [0, 1], *α* = 1 results in LASSO and *α* = 0 in ridge regression, i.e. *L*_2_ regularization. Elastic-net enables a weighted solution that combines the advantages of both LASSO and ridge regression. Similarly to LASSO, elastic-net can also provide sparse and interpretable representations, but in addition it also encourages grouping of regressors, e.g. strongly correlated regressors are typically included or excluded altogether (Zou and Hastie, 2005). However, usage of elastic-net in an optimal manner requires tuning the mixing parameter *α*. Following Occam’s razor principle to create the most simple model that can explain the data, we selected *α* = 1, which is pure LASSO. We train our regression model using the chronological age of each subject as ground truth and estimate the deviances through a 5-fold cross-validation.

#### 2.4.1 Algorithm regressor count

We hypothesize that our dataset is composed of normally aging subjects, i.e., subjects whose chronological age closely matches their brain age, and subjects aging abnormally, i.e., subjects whose chronological age deviates from their brain age. However, we do not know the size of these groups, which subjects belong to each group, their degree of abnormality, nor whether they deviate by aging faster or slower. We first aim to create a model for normal aging subjects; thus, we iteratively exclude outlier subjects. Then, we explore the creation of different models for the different sets of outliers to identify features that could explain abnormal aging.

Iteratively removing outliers from a dataset can potentially be considered a form of p-value hacking in the sense that it reduces the MAE and potentially make the model statistically more significant compared to alternative models. In general, p-value hacking involves manipulating data or analysis procedures in a way that increases the chances of obtaining a significant p-value. In our case, we have a dataset that likely contains outliers in the form of subjects with abnormally aging brains, e.g. due to dementia or any other condition that has not yet impaired function. Outliers are data points that are significantly different from the rest of the data. They can sometimes have a strong influence on statistical analyses, leading to changes in results and interpretations. Removal of outliers should be based on sound statistical or domain-specific reasons, not merely to obtain a desired outcome. Since there is no variable that can be used to identify abnormal aging and remove these outliers, we have to resort to statistical means to remove them. In our study, we clearly document the removal of outliers from the dataset to ensure that their removal is based on objective criteria and not driven by the goal of achieving a specific performance.

We initialize our outlier removal algorithm by introducing a set of kept subjects and outliers. In the first iteration, all subjects belong to the kept set, and none of them are considered outliers. We then proceed to conceptually remove the subject that makes the model deviate the most from the unknown true model. More precisely, we in each iteration employed a Monte-Carlo method to 100 times sample subjects with replacement, i.e. bootstrapping, from the set of kept subjects and fit LASSO models with different regularization coefficients. Each time we selected the model with the minimum deviance and recorded which features were included in that model. After the 100 samples, we trained a linear model on the features, i.e. regressors, that were selected at least once during the Monte-Carlo sampling. We included features in decreasing order of the number of times they were selected in the Monte-Carlo sampling. The model was evaluated using Leave-One-Out Mean Absolute Error (LOOMAE) on the kept subject set. In each iteration, our best model is the one that minimized the LOOMAE. This model with fixed regressors was then retrained on all the subjects, and the residuals were calculated using the retrained model. The subject with the largest residual is appended to the outlier set of subjects and removed from the kept set.

We repeated this process of selecting models based on their regressor count until we could clearly observe that the LOOMAE, as a function of kept subjects, displayed an increasing trend, which happened to be 91 subjects. An increasing trend in LOOMAE means too many subjects have been removed so the model trained on a subset is no longer able to predict the one left out. Thus the best model exist in the valley of the LOOMAE. Due to random noise in the data some models are overfit, i.e. explain noise, and other are underfit, i.e. does not explain the data. Both generalises more poorly to new data than a model that is neither over- nor underfit. To minimise the risk of selecting an overfit or underfit model, we estimate the expected LOOMAE by fitting a polynomial of degree 4 to the observed values and selected the sparsest model in the valley that has a LOOMAE near the expected value. This final model with 39 features is the one most likely to truly capture normal brain aging and yield predicted brain age close to the chronological age for any subjectt with at normally aging brain. This of course only holds under the assumption that the dataset is informative and holds enough normally aging subjects that the outliers can be removed before the ability to predict left out subjects degrade. We refer to this algorithm as Bootstrapped regressor count, and the pseudocode for this algorithm is described in the Appendix.

This model can accurately predict the chronological age of normally aging subjects. The outliers are subjects that have shown irregular signals and whose age cannot be modeled accurately with the same regressors as the kept subjects. We divided the outliers of the selected model into the four cohorts: younger predicted to be older, younger predicted to be younger, older predicted to be younger, and older predicted to be older. Younger subjects are those whose real age is below the mean of 48.04 years, and likewise, older subjects are those whose age is above the mean. We then decided to further explore which features are relevant to better predict these abnormally aging subjects by iteratively reintegrating features into the selected model.

#### 2.4.2 Reintegration of features

To identify which regions were relevant for predicting chronological age, we decided to iteratively reintegrate features into each of the four outlier groups, as shown in Figure 1. We started with the model of the kept subjects with the 39 features as our base model. In each reintegration step, we iteratively selected one feature from all other features not included in the initial 39 features and added it to the set of features of the base model. The model was then refitted using the cohort of subjects, and the LOOMAE was calculated. When refitting the model with the added feature, the parameters of the base model were not updated, and only the weight associated with the reintegrated feature was optimized. The feature resulting in the lowest LOOMAE was reintegrated into the model, and this model became the base model for the next iteration. We continued the procedure until the LOOMAE was equal to or less than the LOOMAE of the model with the 108 kept subjects, which was 2.487 years.

**Figure 1:**
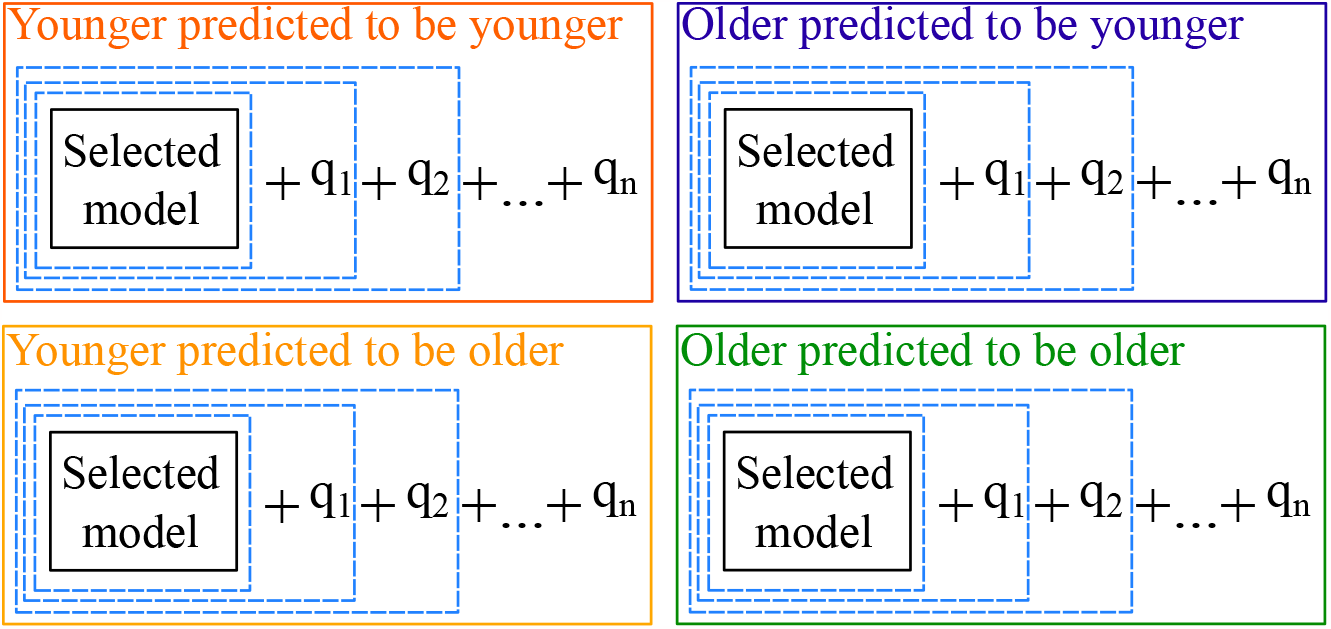
Reintegration of features paradigm. For each of the four abnormally ageing cohorts we reintegrated *n* features until the LOOMAE of the set was lower than the LOOMAE of the models for the normally ageing subjects of 2.487 years. Each reintegrated feature is the feature that minimized the LOOMAE for that iteration. The blue box represents that at each iteration the base model adds one feature and its parameters become fixed before integrating the next feature.

For each cohort, the reintegrated features allowed us to generate hypotheses about which features are relevant to explain abnormal aging. This is because each cohort represents abnormally aging subjects whose brain age cannot be explained with the same model as the majority of other subjects. The reintegrated features were then mapped back to their corresponding brain subnetworks. Table 4 shows the subnetworks considered in this study along with their corresponding brain regions.

**Table 4:** Mapping of the brain features to their respective subnetwork. Indices correspond to the region numbers which range from 1 to 264. **Abbreviations**: *Motor:* Motor network, *CON:* Cingulo-Opercular Network, *Aud:* Auditory network, *DMN:* Default Mode Network, *Vis:* Yisual network, *FPN:* Fronto-Parietal Network, *SAN:* Salience Network, *Subc:* Subcortical Network, *VAN:* Yentral Attention Network, *DAN:* Dorsal Attention Network.

| Subnetwork | Indices | No. regions |
| --- | --- | --- |
| Motor | 13-41, 255 | 29 |
| CON | 47-60 | 13 |
| Aud | 61-73 | 12 |
| DMN | 74-83, 86-131, 137, 139 | 56 |
| Vis | 143-173 | 30 |
| FPN | 174-181, 186-202 | 23 |
| SAN | 203-220 | 17 |
| Subc | 222-234 | 12 |
| VAN | 235-242 | 7 |
| DAN | 251-252, 256-264 | 10 |
| Others |  | 55 |

#### 2.4.3 Monte Carlo simulations for hypothesis testing

Testing of the hypothesis that the observed number of features involving DMN is due to chance was done through a Monte Carlo simulation. In each of the 10,000 iterations, the number of additional features was drawn from a uniform distribution of all possible features without replacement, assuming an equal probability of selecting any one feature. This simulation was done in Python 3 using the NumPy package.

## 3 Results

### 3.1 Model selection

Figure 2 displays the LOOMAE error as a function of the number of subjects for the bootstrapped regressor count algorithm. The LOOMAE curve exhibits a parabolic shape, which is expected since, at the beginning, the dataset contains all outliers that negatively impact the performance of the models. As we iteratively remove outliers, the LOOMAE of the models decreases and converges to a low level, as evidenced by the LOOMAE from approximately 100 to 130 subjects being, on average, around 2.5 years. Subsequently, removing too many subjects causes the LOOMAE of the models to increase again, as the information in the remaining small number of subjects is insufficient to explain each other, unlike when it converged to the best solutions. Note that the LOOMAE fluctuates around the expected value due to the inclusion/exclusion of features needed to explain random noise in the data. We fit a degree-4 polynomial to the LOOMAE to obtain an estimate of the expected LOOMAE. This polynomial has a global minimum of 110.49 subjects. To minimize the risk of over-/underfitting to random noise, we choose to work with a model that has the LOOMAE close to the expected one. This model with 108 subjects also strikes a good balance between LOOMAE and the number of included features, having only 39 features. Models with LOOMAE values below the fitted line are overfitted, as the fitted curve represents the expected LOOMAE for that specific number of subjects.

**Figure 2:**
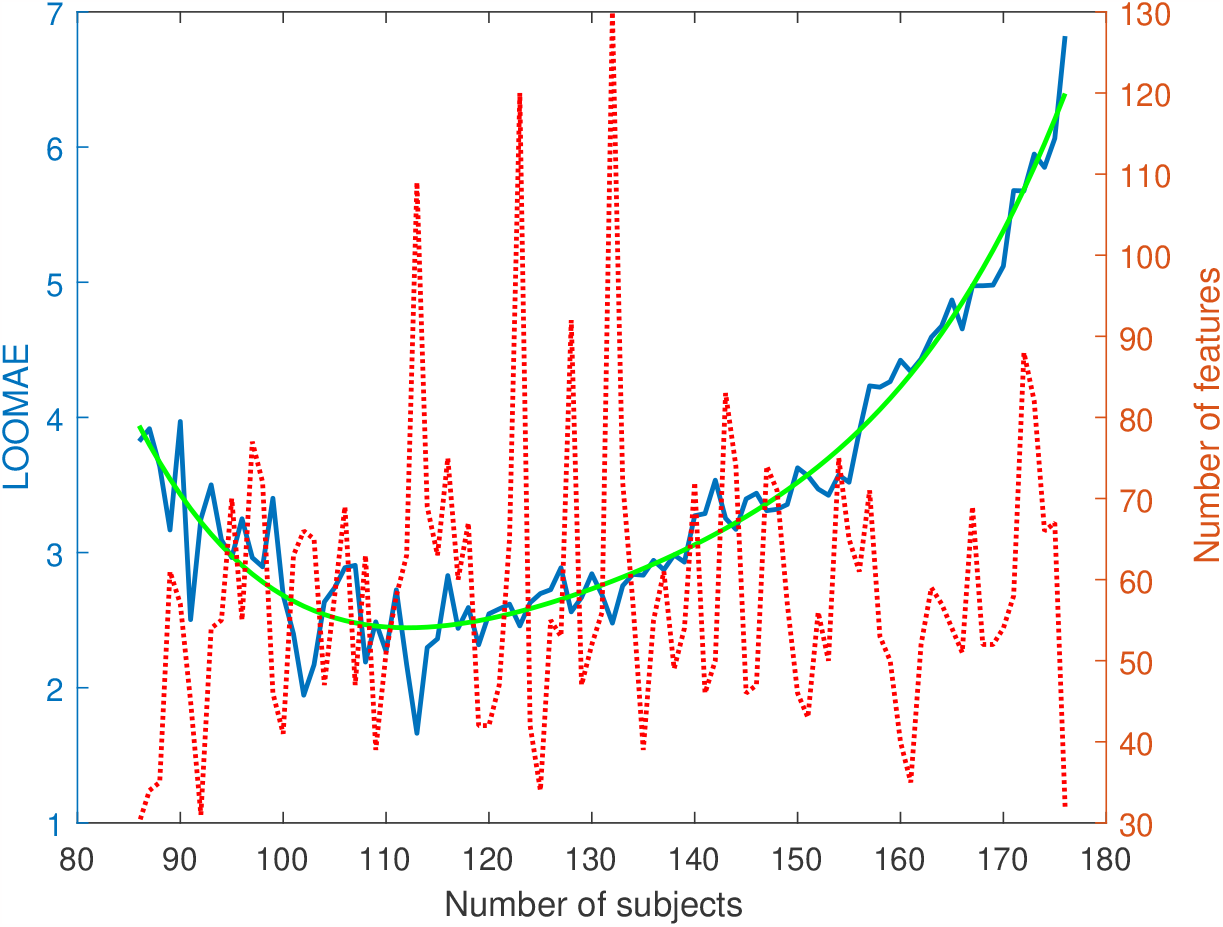
Number of kept subjects vs LOOMAE of selected model (blue solid line), fitted polynomial curve of degree 4 (green solid line), and the number of features of the selected model for that iteration (red dotted line).

We chose this model because the LOOMAE agrees with the expected one, the number of features is low relative to the total number of subjects, and it is close to the minimum of the expected LOOMAE. At the minimum, due to noise, we happened to get models that have around twice the number of features compared to the selected model and are clearly overfitted to the selected dataset, and therefore poor choices. The demographic information of the 68 removed subjects is included in the Appendix.

Figure 3 shows the error distribution for the model with 108 subjects organized by decades. The model tends to predict subjects with chronological age below the mean of 48.04 years as older and subjects with chronological age above the mean as younger. This makes sense as the model is simply a linear combination of the 39 selected features and lacks the complexity to capture the nuisances at the extremes of the distribution.

**Figure 3:**
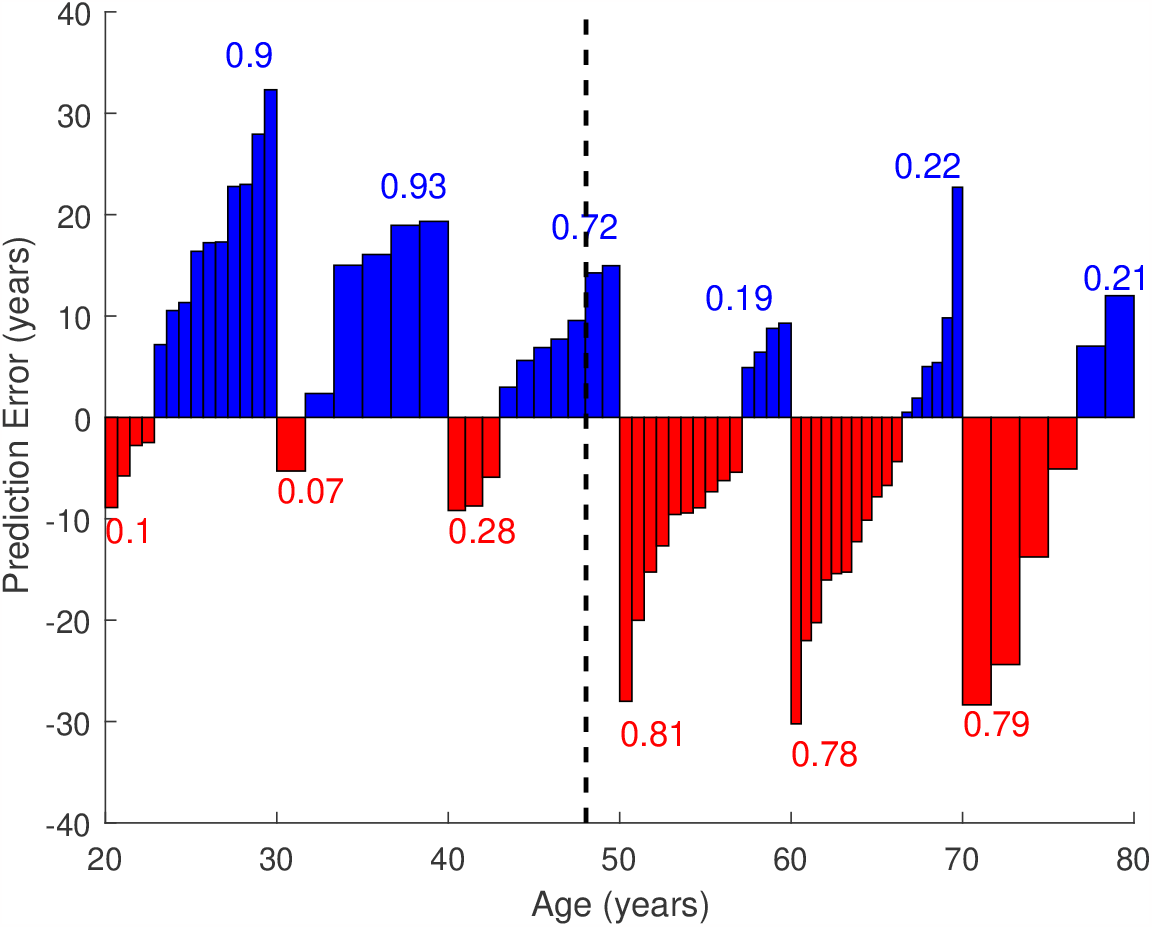
The distribution of the prediction error, *i*.*e*. predicted age minus chronological age, of the outliers using the model with 108 subjects organized by decades. The numbers denote the ratio of the red and blue areas for each decade, i.e. the fraction of subjects predicted to be older and younger, respectively. The dotted black line marks the average chronological age of all subjects (48.04 years).

The number of times a feature was selected and the LOOMAE as a function of the included features is shown in Figure 4. We can see how the number of times a feature was selected exponentially decreases with every feature added. The LOOMAE seems to follow a similar decreasing trend until 39 added features, after which an increase is observed. The selected model had 39 features and a LOOMAE of 2.487 years on the kept subjects. Out of the 68 outliers of this model, 23 are younger subjects predicted to be older (33.8%), s are younger subjects predicted to be younger (11.8%), 25 are older subjects predicted to be younger (36.8%), and 12 are older subjects predicted to be older (17.6%).

**Figure 4:**
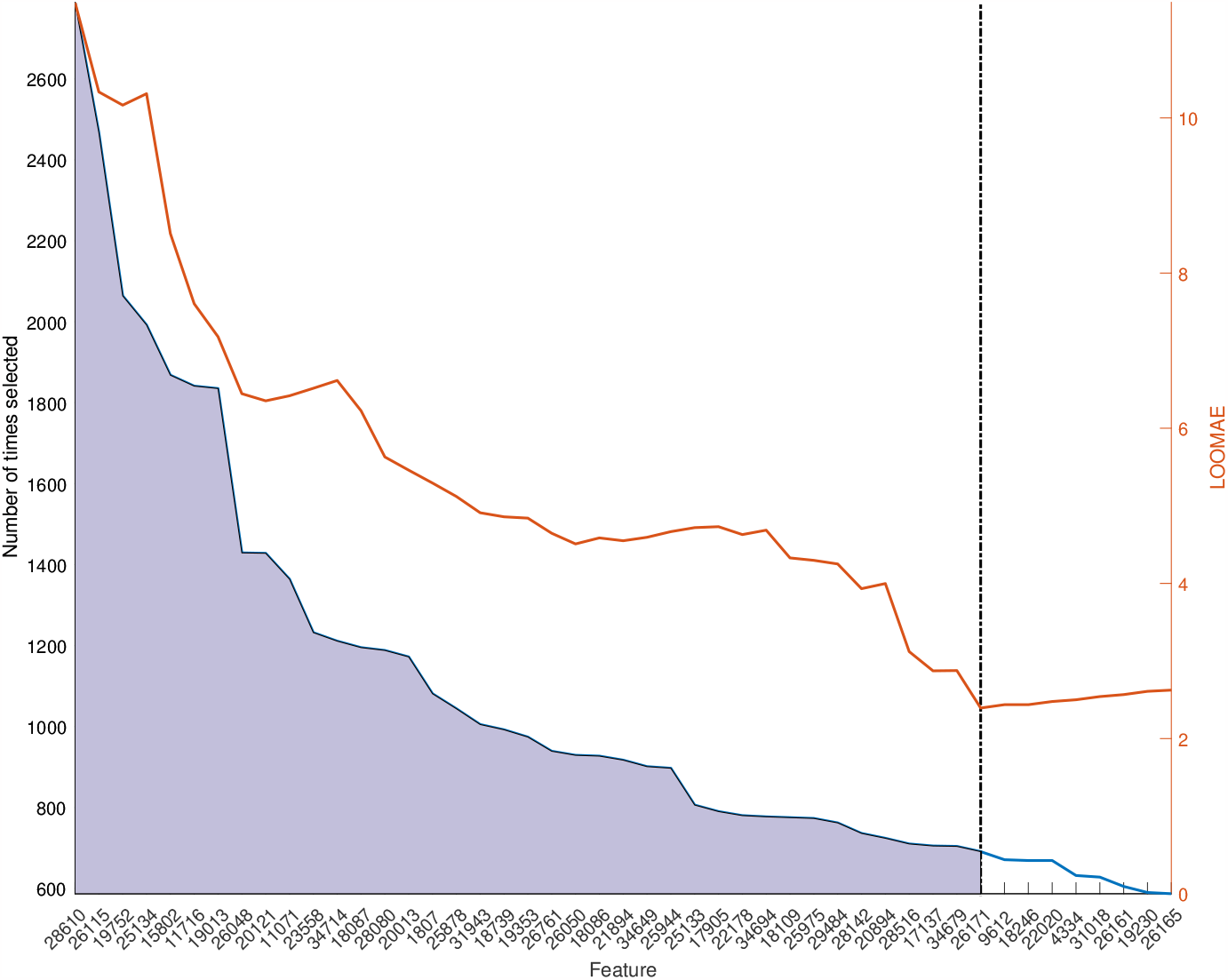
Feature cuts and LOOMAE for the most selected features in the model for 108 subjects. The features are in decreasing order of number of times selected. The selected model for 108 subjects includes the 39 features to the left of the black dotted line. The model yields a leave-one-out absolute error of 2.487 for the kept subjects.

The predicted age as a function of the real age is shown in Figure 5. The model is able to model well the kept subjects with a MAE of 1.58 years and does not perform well on the outliers with a MAE of 12.32 years. In this case, MAE is the average absolute value of the residuals of the model trained on the kept subjects evaluated on all samples of the dataset. It differs from the LOOMAE in the sense that the LOOMAE is calculated by training on all samples except one and evaluating the excluded sample until each sample serves as the left out sample. The LOOMAE then averages all the absolute values of the residuals of left out samples. The accuracy of the model for predicting kept subjects is also represented by the position of the points relative to the red dashed line, which represents the perfect model that always predicts the brain age the same as the chronological age. It is also noticeable that the model performs equally well on the kept subjects, independent of their age, while overestimating the age of younger outliers and underestimating the age of older outliers.

**Figure 5:**
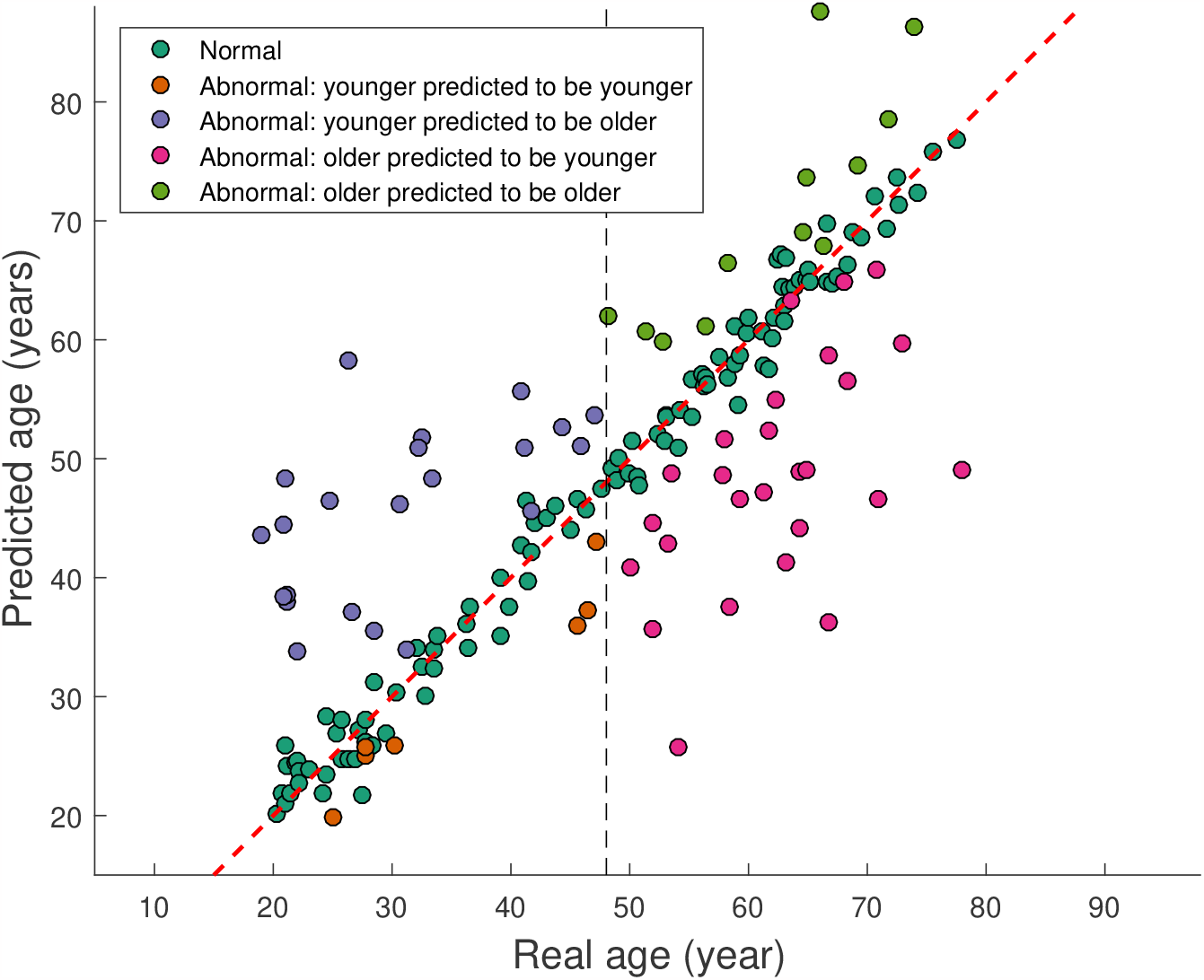
Predicted age as a function of real age for the model of 108 subjects for the normally ageing subjects (blue) and the four abnormally ageing cohorts of younger predicted to be younger (orange), younger predicted to be older (yellow), older predicted to be younger (purple) and older predicted to be older (green). The red dashed line marks the *y* = *x* line. Any deviations from the red line represent deviations from the perfect model. The black dashed line marks the mean age of the subjects of 48.04 years.

### 3.2 Reintegration of features

Table 5, Table 6, Table 7, and Table 8 show the results of the reintegration of features for the younger subjects predicted older, younger subjects predicted to be younger, older subjects predicted to be younger, and older subjects predicted to be older, respectively. The mapping of brain regions to their respective subnetworks is depicted in Table 4. We can see that the younger subjects predicted to be older and the older subjects predicted to be younger have the largest LOOMAE values of 14.974 and 14.198 years, respectively. These are the two cohorts with the largest number of outliersâ€”23 and 25, respectively. The younger subjects predicted to be younger and the older subjects predicted to be older have LOOMAE of 6.129 and 7.818 years and only contain s and 12 years, respectively.

**Table 5:** Best model (model for 108 subjects) analysis for the Younger subjects predicted to be older. Age^*n*^ is the predicted age after adding *n* features. **Feature No**. refers to the index of the feature in our dataset which ranges from 1 to 34716, **Region No**. refers to the brain regions which is a tuple in which each element ranges from 1 to 264, **Region name** is the name of the subnetwork associated with each of the 264 regions; in case there was no subnetwork associated with a region we report the **Region No**..

|  | Subject ID | Real age | Predicted age | Age<sup>1</sup> | Age<sup>2</sup> | Age<sup>3</sup> | Age<sup>4</sup> |
| --- | --- | --- | --- | --- | --- | --- | --- |
|  | 138 | 20.986 | 48.925 | 15.809 | 24.869 | 17.624 | 23.769 |
|  | 191 | 26.279 | 58.597 | 11.685 | 35.344 | 19.184 | 32.724 |
|  | 145 | 21.148 | 37.534 | 21.818 | 25.424 | 20.214 | 21.989 |
|  | 148 | 32.523 | 51.862 | 35.604 | 30.718 | 33.449 | 33.054 |
|  | 173 | 30.578 | 46.646 | 28.261 | 25.61 | 34.214 | 29.099 |
|  | 151 | 21.156 | 38.388 | 24.785 | 13.794 | 25.533 | 17.114 |
|  | 189 | 18.995 | 43.7 | 8.267 | 24.58 | 13.427 | 22.79 |
|  | 102 | 22.033 | 33.363 | 26.144 | 18.68 | 22.255 | 23.326 |
|  | 83 | 48.14 | 62.393 | 54.972 | 46.156 | 44.997 | 47.848 |
|  | 174 | 33.364 | 48.375 | 31.722 | 35.459 | 34.377 | 33.374 |
|  | 146 | 20.863 | 38.166 | 19.72 | 15.045 | 27.975 | 13.579 |
|  | 178 | 44.332 | 52.054 | 45.68 | 47.344 | 41.963 | 45.402 |
|  | 74 | 40.814 | 55.778 | 40.244 | 39.863 | 42.339 | 38.866 |
|  | 142 | 24.682 | 47.665 | 17.998 | 30.676 | 24.038 | 24.651 |
|  | 176 | 32.255 | 51.204 | 35.348 | 27.588 | 33.785 | 32.35 |
|  | 133 | 20.836 | 43.615 | 18.062 | 24.283 | 20.889 | 19.283 |
|  | 160 | 26.668 | 37.208 | 23.21 | 27.058 | 23.844 | 26.460 |
|  | 164 | 28.523 | 35.702 | 33.825 | 28.009 | 32.048 | 29.250 |
|  | 82 | 41.129 | 50.687 | 40.357 | 41.237 | 41.323 | 40.483 |
|  | 96 | 45.94 | 51.556 | 45.935 | 45.297 | 45.651 | 44.437 |
|  | 77 | 47.093 | 53.982 | 47.458 | 46.958 | 47.579 | 47.212 |
|  | 67 | 41.715 | 44.698 | 51.089 | 40.192 | 40.336 | 44.863 |
|  | 165 | 31.216 | 33.575 | 43.275 | 27.084 | 34.225 | 29.346 |
| Feature No. |  |  |  | 5570 | 20376 | 21677 | 23537 |
| Region No. |  |  |  | (38,23) | (120,95) | (105,103) | (114,260) |
| Region name |  |  |  | Motor-Motor | DMN-DMN | DMN-DMN | DMN-DAN |
| LOOMAE |  |  | 14.974 | 4.680 | 3.661 | 2.612 | 1.950 |

**Table 6:** Best model (model for 108 subjects) analysis for the Younger subjects predicted to be younger. Age^*n*^ is the predicted age after adding *n* features. **Feature No**. refers to the index of the feature in our dataset which ranges from 1 to 34716, **Region No**. refers to the brain regions which is a tuple in which each element ranges from 1 to 264, **Region name** is the name of the subnetwork associated with each of the 264 regions; in case there was no subnetwork associated with a region we report the **Region No**..

|  | Subject ID | Real age | Predicted age | Age<sup>1</sup> |
| --- | --- | --- | --- | --- |
|  | 91 | 45.584 | 36.401 | 45.954 |
|  | 171 | 30.249 | 24.959 | 30.935 |
|  | 153 | 24.992 | 19.216 | 25.219 |
|  | 10 | 47.197 | 41.297 | 46.794 |
|  | 149 | 27.775 | 25.005 | 26.9 |
|  | 194 | 27.762 | 25.279 | 28.044 |
|  | 86 | 46.485 | 37.747 | 46.251 |
|  | 167 | 27.444 | 18.552 | 27.391 |
| Feature No. |  |  |  | 1903 |
| Region No. |  |  |  | (91,8) |
| Region name |  |  |  | DMN-8 |
| LOOMAE |  |  | 6.129 | 0.520 |

**Table 7:** Best model (model for 108 subjects) analysis for the Older subjects predicted to be younger. Age^*n*^ is the predicted age after adding *n* features. **Feature No**. refers to the index of the feature in our dataset which ranges from 1 to 34716, **Region No**. refers to the brain regions which is a tuple in which each element ranges from 1 to 264, **Region name** is the name of the subnetwork associated with each of the 264 regions; in case there was no subnetwork associated with a region we report the **Region No**..

|  | Subject ID | Real age | Predicted age | Age<sup>1</sup> | Age<sup>2</sup> | Age<sup>3</sup> |
| --- | --- | --- | --- | --- | --- | --- |
|  | 11 | 63.216 | 41.2 | 69.657 | 59.269 | 65.194 |
|  | 197 | 64.332 | 44.085 | 64.905 | 68.149 | 63.087 |
|  | 203 | 66.688 | 36.46 | 81.676 | 55.702 | 71.252 |
|  | 188 | 59.2 | 46.53 | 58.183 | 57.322 | 59.842 |
|  | 112 | 54.132 | 26.126 | 62.896 | 52.891 | 52.713 |
|  | 14 | 70.879 | 46.5 | 77.189 | 64.861 | 73.193 |
|  | 60 | 77.992 | 49.638 | 83.117 | 73.105 | 78.728 |
|  | 17 | 53.238 | 43.81 | 47.726 | 55.19 | 52.626 |
|  | 117 | 57.808 | 48.888 | 56.757 | 59.747 | 56.928 |
|  | 62 | 57.986 | 51.748 | 48.282 | 58.61 | 57.402 |
|  | 47 | 68.329 | 56.067 | 73.234 | 67.705 | 68.826 |
|  | 8 | 66.677 | 58.845 | 69.155 | 64.292 | 71.918 |
|  | 50 | 51.926 | 36.67 | 49.014 | 56.46 | 52.335 |
|  | 42 | 61.299 | 46.034 | 56.546 | 63.83 | 61.424 |
|  | 33 | 64.236 | 48.199 | 66.005 | 62.543 | 64.383 |
|  | 192 | 61.715 | 51.577 | 56.313 | 61.224 | 66.93 |
|  | 66 | 53.46 | 48.049 | 50.618 | 54.703 | 55.077 |
|  | 71 | 50.077 | 40.497 | 49.402 | 51.227 | 48.912 |
|  | 119 | 58.438 | 38.424 | 58.217 | 62.317 | 55.895 |
|  | 75 | 51.937 | 44.603 | 51.741 | 54.983 | 49.308 |
|  | 5 | 64.811 | 49.403 | 65.001 | 67.827 | 63.296 |
|  | 29 | 62.247 | 55.532 | 64.717 | 65.551 | 58.99 |
|  | 63 | 70.718 | 65.631 | 58.549 | 77.664 | 67.394 |
|  | 185 | 67.978 | 63.619 | 60.453 | 64.439 | 65.363 |
|  | 26 | 72.893 | 59.126 | 72.859 | 72.599 | 71.194 |
| Feature No. |  |  |  | 19125 | 30276 | 2169 |
| Region No. |  |  |  | (249,87) | (195,170) | (102,9) |
| Region name |  |  |  | 249-DMN | FPN-Vis | DMN-9 |
| LOOMAE |  |  | 14.198 | 4.661 | 3.305 | 2.090 |

**Table 8:** Best model (model for 108 subjects) analysis for the Older subjects predicted to be older. Age^*n*^ is the predicted age after adding *n* features. **Feature No**. refers to the index of the feature in our dataset which ranges from 1 to 34716, **Region No**. refers to the brain regions which is a tuple in which each element ranges from 1 to 264, **Region name** is the name of the subnetwork associated with each of the 264 regions; in case there was no subnetwork associated with a region we report the **Region No**..

|  | Subject ID | Real age | Predicted age | Age<sup>1</sup> | Age<sup>2</sup> |
| --- | --- | --- | --- | --- | --- |
|  | 41 | 66.058 | 88.759 | 60.62 | 68.699 |
|  | 24 | 66.274 | 68.174 | 66.096 | 65.739 |
|  | 21 | 69.233 | 74.249 | 72.53 | 65.51 |
|  | 13 | 63.526 | 64.032 | 63.254 | 62.759 |
|  | 53 | 52.827 | 59.26 | 58.86 | 52.464 |
|  | 36 | 73.874 | 85.879 | 74.39 | 74.024 |
|  | 68 | 64.66 | 70.07 | 62.474 | 66.622 |
|  | 52 | 56.444 | 61.36 | 58.612 | 57.918 |
|  | 126 | 64.907 | 74.724 | 64.768 | 64.454 |
|  | 115 | 51.353 | 60.135 | 51.226 | 51.477 |
|  | 84 | 58.244 | 67.536 | 57.107 | 57.108 |
|  | 37 | 71.734 | 78.768 | 69.197 | 72.361 |
| Feature No. |  |  |  | 26482 | 1638 |
| Region No. |  |  |  | (158,136) | (82,7) |
| Region name |  |  |  | Vis-136 | DMN-7 |
| LOOMAE |  |  | 7.818 | 2.571 | 1.400 |

Among these four cohorts, the largest number of additional features to reduce the LOOMAE to the target LOOMAE of 2.487 years was four for the younger predicted older. Of these four features, three involve the DMN (75%). The older predicted to be younger required three additional features to reduce the LOOMAE below the target, of which two involve the DMN (67%). The older predicted to be older required two additional features to reduce the LOOMAE below the target, of which one involves the DMN (50%). Even the younger predicted to be younger cohort, which required only one additional feature, included a feature associated with the DMN (100%). We find it interesting that features involving the DMN were added in all four cases and investigate the likelihood of this.

Of the 39 features in our model for predicting brain age in normally aging subjects 17 (43.59%) involve the DMN, which is lower than the incidence among the additional features in the four cohorts. Given that 56 out of 264 regions corresponding to the DMN (21.21%), as shown in Table 4, the proportion of the 34 716 features involving DMN is 37.99%, since each feature is a correlation involving two of the 264 brain regions. The expected number of 39 randomly picked features involving DMN is only 14.8 and the probability of at random picking 17 or more is 0.29. The expected number of 4 randomly picked features involving DMN is only 1.5 and the probability of at random picking 3 or more is 0.16. The expected number of randomly picked features involving DMN is only 1.2 and the probability of at random picking 2 or more is 0.32. The expected number of 2 randomly picked features involving DMN is 0.8 and the probability of at random picking 1 or more is 0.62. The expected number of 1 randomly picked features involving DMN is 0.38 and the probability of at random picking 1 or more is 0.38. In total 7 out of the 10 additional features involves DMN. The expected number is 3.8 and the probability of at random picking 7 or more is 0.035. While the enrichment of DMN in any of the sets of additional features is not statistically significant, it is when considering all four sets. The hypothesis that the observed number of features involving DMN is due to chance is rejected, assuming equal probability of selecting any one feature. For this reason, we believe that the DMN is an essential feature to predict the brain age of abnormally aging subjects. We cannot rule out that this enrichment of DMN is due to our usage of rsfMRI and the DMN therefore being active.

## 4 Conclusions

Our results indicate that we have successfully created a model to predict the age of normally aging subjects by removing outliers with abnormally aging brains. This model has the lowest prediction error among the models used to predict age over a lifespan based on neuroimaging data, in terms of MAE. Having a model that accurately predicts age for normally aging subjects enables researchers to concentrate on characterizing the natural aging trajectory without the influence of outliers or confounding factors associated with abnormal aging. The model can also serve as a screening tool, as it can effectively identify subjects whose biological age significantly differs from their chronological age, potentially indicating underlying health issues or neurodegenerative conditions that require further investigation.

We also identified regions that could explain abnormal aging with the model for normally aging subjects as a base. Among these, the DMN revealed itself to be relevant in all of the abnormal cohorts. This subnetwork is highly active during rest and becomes less active during goal-directed cognitive tasks. It is believed to play a crucial role in various cognitive functions, including self-referential thinking, social cognition, and memory consolidation. Our models are consistent with neuroscience theories related to brain aging and network dynamics.

We believe our contribution plants a seed towards enabling timely interventions to slow or prevent neurodegenerative diseases. Additionally, our identification of the DMN as a relevant brain region for abnormal aging may serve as a biomarker for certain conditions and therefore improve diagnostics and personalized treatments. The interpretability of our model allows for easy understanding by most stakeholders of the connectivity patterns associated with aging. Prioritizing the identified brain regions can lead to initiatives for the early detection of abnormalities, which can further improve brain health in aging populations.

## 5 Author contributions

Author contribution using the CRediT taxonomy: Conceptualization: SH and TN; Methodology: JC and TN; Software: JC and ZY; Yalidation: JC and TN; Formal analysis: JC; Investigation: JC; Resources: SH and TN; Data curation: SH and ZY; Writing - original draft preparation: JC; Writing - review and editing: SH, TN, and ZY; Yisualization: JC and TN; Supervision: SH and TN. Project administration: SH and TN; Funding acquisition: SH and TN;

## 7 Data availability

The data used in this study is not publicly available due to ethical and privacy considerations. The data utilized for our research is the property of the Mind Research Imaging Center at National Cheng Kung University. Access to the data may be granted in accordance with the ethical and legal guidelines established by the institution. For inquiries or requests regarding data access, interested parties should contact the Mind Research Imaging Center at National Cheng Kung University for further information and permissions.

## 8 Acknowledgments

We thank the Mind Research and Imaging Center (MRIC), supported by MOST, at NCKU for consultation and instrument availability.

## A Appendix

### A.1 Feature selection using LASSO

Suppose we have a dataset of a *n* number of *p*-dimensional observations where *p >> n*. Each observation consists of a scalar target *y* and an input vector *x* ∈ *Iℝ*^*p*^. Linear models, in the form of

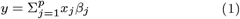

where *β*_*j*_ are the parameters of the model, serve as a method for fitting the data and describing the relation between the input variables *x* and the target *y*. In this case, the parameters *β*_*j*_ describe how much weight or relevance each variable has on the output.

Ordinary least squares (OLS) is a technique for estimating the optimal parameters in a linear regression model. The values of the parameters are selected through the principle of least squares, *i*.*e*. minimizing the sum of the residuals squared between the target *y* and the prediction of the linear model. In more general terms, the objective function of OLS can be written as

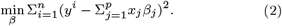

LASSO formulates the problem of estimating the model parameters *β*_*j*_ as

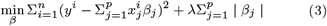

where *λ >* 0 is the *L*1 regularization parameter that defines the amount of parameters in solution *β* (Huang and Jojic, 2011). *The value of λ* influences the sparsity of the model as a higher *λ* yields a sparser model.

### A.2 Outlier selection algorithm

We include the pseudocode of our algorithm used to select outliers which we coined bootstrap regressor count. The list of outliers in order of removal for our selected model can be seen on Table 9.

**Table 9:** Outliers information table for 68 removed subjects. The outliers are sorted from first removed (top) to last removed (bottom). For Gender, 0 means male. For MRI update, 0 means before the update. For Cardiovascular risk factors, 1 indicates that the participant has at least one risk factor.

| Original ID | Gender | Education years | MoCA | BDI | Cardiovascular risk factor | MRI update | Real age | Predicted age | Error |
| --- | --- | --- | --- | --- | --- | --- | --- | --- | --- |
| 138 | 0 | 15 | 28 | 1 | 0 | 0 | 20.986 | 48.925 | 27.938 |
| 11 | 0 | 12 | 28 | 7 | 0 | 0 | 63.216 | 41.2 | -22.017 |
| 191 | 0 | 13 | 30 |  |  | 1 | 26.279 | 58.597 | 32.318 |
| 145 | 1 | 15 | 29 | 2 | 0 | 0 | 21.148 | 37.534 | 16.386 |
| 148 | 0 | 16 | 28 | 1 | 0 | 0 | 32.523 | 51.862 | 19.339 |
| 197 | 0 | 16 | 30 |  |  | 0 | 64.332 | 44.085 | -20.247 |
| 203 | 0 | 16 | 28 |  |  | 0 | 66.688 | 36.46 | -30.227 |
| 188 | 1 | 16 | 30 |  |  | 0 | 59.2 | 46.53 | -12.67 |
| 112 | 1 | 16 | 26 | 0 | 0 | 0 | 54.132 | 26.126 | -28.005 |
| 173 | 0 | 18 | 29 | 4 | 0 | 0 | 30.578 | 46.646 | 16.068 |
| 14 | 0 | 12 | 26 | 5 | 0 | 0 | 70.879 | 46.5 | -24.38 |
| 151 | 1 | 15 | 29 | 7 | 0 | 0 | 21.156 | 38.388 | 17.232 |
| 60 | 0 | 12 | 26 | 4 | 0 | 0 | 77.992 | 49.638 | -28.353 |
| 189 | 1 | 13 | 29 |  |  | 1 | 18.995 | 43.7 | 24.706 |
| 41 | 0 | 16 | 25 | 10 | 1 | 0 | 66.058 | 88.759 | 22.701 |
| 102 | 1 | 16 | 30 | 4 | 0 | 0 | 22.033 | 33.363 | 11.33 |
| 17 | 1 | 12 | 28 | 0 | 0 | 0 | 53.238 | 43.81 | -9.4287 |
| 24 | 0 | 16 | 27 | 0 | 0 | 0 | 66.274 | 68.174 | 1.9 |
| 83 | 1 | 12 | 29 | 5 | 1 | 0 | 48.14 | 62.393 | 14.253 |
| 21 | 0 | 12 | 27 | 6 | 1 | 0 | 69.233 | 74.249 | 5.0159 |
| 117 | 1 | 16 | 27 | 0 | 1 | 0 | 57.808 | 48.888 | -8.9199 |
| 91 | 0 | 14 | 28 | 3 | 1 | 0 | 45.584 | 36.401 | -9.1821 |
| 13 | 1 | 12 | 25 | 2 | 0 | 0 | 63.526 | 64.032 | 0.50611 |
| 62 | 0 | 16 | 28 | 2 | 0 | 0 | 57.986 | 51.748 | -6.2379 |
| 174 | 0 | 16 | 26 | 13 | 0 | 0 | 33.364 | 48.375 | 15.011 |
| 47 | 1 | 14 | 27 | 2 | 0 | 0 | 68.329 | 56.067 | -12.262 |
| 53 | 0 | 18 | 27 | 2 | 0 | 0 | 52.827 | 59.26 | 6.4328 |
| 146 | 0 | 15 | 28 | 3 | 0 | 0 | 20.863 | 38.166 | 17.303 |
| 178 | 0 | 14 | 24 | 0 | 0 | 0 | 44.332 | 52.054 | 7.7228 |
| 8 | 1 | 14 | 27 | 13 | 0 | 0 | 66.677 | 58.845 | -7.8316 |
| 74 | 0 | 14 | 23 | 2 | 0 | 0 | 40.814 | 55.778 | 14.964 |
| 142 | 1 | 16 | 29 | 9 | 0 | 0 | 24.682 | 47.665 | 22.983 |
| 171 | 0 | 16 | 27 | 12 | 0 | 0 | 30.249 | 24.959 | -5.2901 |
| 36 | 0 | 18 | 27 | 1 | 0 | 0 | 73.874 | 85.879 | 12.005 |
| 50 | 1 | 14 | 27 | 9 | 0 | 0 | 51.926 | 36.67 | -15.256 |
| 42 | 1 | 16 | 25 | 3 | 0 | 0 | 61.299 | 46.034 | -15.264 |
| 176 | 1 | 14 | 28 | 7 | 0 | 0 | 32.255 | 51.204 | 18.949 |
| 68 | 0 | 15 | 26 | 0 | 0 | 0 | 64.66 | 70.07 | 5.4102 |
| 153 | 0 | 16 | 29 | 0 | 0 | 0 | 24.992 | 19.216 | -5.7753 |
| 10 | 0 | 12 | 24 | 7 | 0 | 0 | 47.197 | 41.297 | -5.8998 |
| 52 | 1 | 16 | 24 | 0 | 0 | 0 | 56.444 | 61.36 | 4.9158 |
| 133 | 0 | 15 | 28 | 4 | 0 | 0 | 20.836 | 43.615 | 22.779 |
| 149 | 0 | 19 | 29 | 8 | 0 | 0 | 27.775 | 25.005 | -2.7708 |
| 160 | 0 | 16 | 27 | 7 | 0 | 0 | 26.668 | 37.208 | 10.539 |
| 164 | 0 | 16 | 28 | 1 | 0 | 0 | 28.523 | 35.702 | 7.1789 |
| 82 | 1 | 16 | 28 | 3 | 0 | 0 | 41.129 | 50.687 | 9.5583 |
| 33 | 1 | 16 | 27 | 3 | 0 | 0 | 64.236 | 48.199 | -16.036 |
| 126 | 1 | 16 | 29 | 4 | 0 | 0 | 64.907 | 74.724 | 9.8172 |
| 192 | 0 | 12 | 29 |  |  | 1 | 61.715 | 51.577 | -10.138 |
| 66 | 1 | 14 | 28 | 6 | 0 | 0 | 53.46 | 48.049 | -5.4116 |
| 115 | 1 | 23 | 26 | 3 | 1 | 0 | 51.353 | 60.135 | 8.7817 |
| 71 | 1 | 12 | 29 | 3 | 0 | 0 | 50.077 | 40.497 | -9.5797 |
| 119 | 1 | 12 | 28 | 0 | 0 | 0 | 58.438 | 38.424 | -20.015 |
| 75 | 1 | 12 | 26 | 7 | 1 | 0 | 51.937 | 44.603 | -7.3337 |
| 194 | 0 | 16 | 29 |  |  | 1 | 27.762 | 25.279 | -2.4825 |
| 96 | 1 | 12 | 25 | 9 | 0 | 0 | 45.94 | 51.556 | 5.6168 |
| 5 | 1 | 11 | 29 | 1 | 1 | 0 | 64.811 | 49.403 | -15.408 |
| 86 | 1 | 14 | 27 | 2 | 0 | 0 | 46.485 | 37.747 | -8.7384 |
| 29 | 1 | 14 | 25 | 10 | 0 | 0 | 62.247 | 55.532 | -6.7144 |
| 77 | 1 | 14 | 29 | 2 | 0 | 0 | 47.093 | 53.982 | 6.8885 |
| 67 | 0 | 14 | 26 | 13 | 0 | 0 | 41.715 | 44.698 | 2.9826 |
| 84 | 0 | 12 | 24 | 8 | 1 | 0 | 58.244 | 67.536 | 9.2924 |
| 63 | 1 | 9 | 23 | 2 | 0 | 0 | 70.718 | 65.631 | -5.0869 |
| 165 | 1 | 18 | 29 | 10 | 0 | 0 | 31.216 | 33.575 | 2.3586 |
| 185 | 0 | 6 | 27 |  |  | 1 | 67.978 | 63.619 | -4.3595 |
| 37 | 0 | 12 | 26 | 0 | 0 | 0 | 71.734 | 78.768 | 7.0339 |
| 26 | 0 | 16 | 28 | 0 | 0 | 0 | 72.893 | 59.126 | -13.767 |
| 167 | 1 | 16 | 28 | 5 | 0 | 0 | 27.444 | 18.552 | -8.8918 |

